# Factor Analysing Predictive Processing: No Evidence for a General Factor Across Tasks

**DOI:** 10.64898/2026.06.09.26354804

**Authors:** Chantal Miller-Silva, Franziska Knolle, Andrea Greve, Franciska de Beer, Tamara Mujirishvili, Lucy J MacGregor, Philip R Corlett, Joost Haarsma, Albert R Powers, Graham K Murray

## Abstract

**Background & Hypothesis:** Dysfunctional predictive processing (PP), specifically the aberrant weighting of priors, is a frequently-proposed mechanism for psychosis and psychosis-like phenomena (schizotypy). Evidence for this theory mostly originates from single-task studies, which assume that all tasks load onto a single latent construct of PP performance, but the underlying factor structure of PP tasks is unknown. PP deficits in psychosis may be better described by a two-factor, hierarchical model: weakened lower-level (perceptual) priors compensated by higher-level (cognitive) priors.

**Study Design:** This study implements a multi-paradigm approach in healthy participants to investigate latent constructs underlying PP and their relationship to schizotypy. Participants (*N* = 73) completed 6 tasks measuring reliance on priors across language, memory, visual, and auditory domains. A factor analysis investigated whether performance across tasks is captured by a single or two-factor model.

**Study Results:** Although a two-factor model best described performance, factors reflected within-task correlations rather than a PP hierarchy. Cross-task PP measures were poorly correlated, suggesting that individuals’ weighting of priors was task-specific. A full model including all task outcomes (not factors) significantly predicted the severity of schizotypal aberrant beliefs but no other schizotypal measures.

**Conclusions:** These results do not evidence a single factor underpinning PP performance. It is therefore inappropriate to use results from single tasks to propose a generalised PP deficit in psychosis. Variation was also not captured by a two-factor hierarchical model of priors. Further multi-paradigm research is required to evaluate alternative models or additional variables that describe aberrant PP in psychosis.

## 1 Introduction

### 1.1 Can a Global Predictive Processing Deficit Explain Psychosis?

Predictive processing is a highly influential theory of brain function, as demonstrated by its applications in sensorimotor,^1^ memory,^2^ language,^3^ auditory,^4^ visual,^5^ and developmental^6^ research. This theory posits that internal models of the world (prior expectations; *priors*) are used to predict incoming sensory signals in a process of prediction error (PE) minimisation that is akin to Bayesian inference.^7^ Priors facilitate faster, more efficient processing of ambiguous stimuli than is achievable with a purely bottom-up strategy, but the ability of priors to colour perception can lead to maladaptive inferences such as illusions.^8^ As such, the aberrant weighting (or ‘strength’) of priors has become a popular theory of psychosis and related subclinical phenomena (*schizotypy*).^9,10^ However, results have been conflicting: different studies report an increased or decreased reliance on priors (i.e., ‘stronger’ and ‘weaker’ priors, respectively) in psychosis and schizotypy,^11–14^ while others indicate intact^15,16^ priors.

Despite the inconsistent findings, studies typically use results from a single task with a modest sample to propose a generalised predictive processing deficit that should be identifiable across diverse tasks. A recent meta-analysis^17^ suggested that this interpretation is unwarranted, as neither a diagnosis of psychosis nor individual symptoms were related to a single predictive processing dysfunction across paradigms targeting different aspects of perception. These conflicting group-level results could suggest that performance across tasks is not underpinned by a single, domain-general factor reflecting reliance on priors at the level of the individual. An alternative theory proposes that discrepant findings may be reconciled by categorising priors according to their position in the processing hierarchy: weaker lower-level *perceptual* priors may trigger a compensatory increased reliance on higher-level *cognitive* priors in psychosis.^18,19^ However, as most research implements only a single task, there is no empirical evidence that this two-level model can predict an individual’s performance across tasks or explain the conflicting results relating to psychotic and schizotypal symptoms.

### 1.2 The Current Study

Multi-paradigm research is required to understand the latent variables underpinning performance on predictive processing paradigms. Developing a framework for how individuals weight priors across tasks is important for advancing theories on the relationship between predictive processing deficits and psychosis. For example, cross-task consistencies may evidence the two-factor hierarchical model of priors, which could reconcile between-paradigm variations in the relationship between predictive processing and psychotic symptoms. In this model, an individual’s reliance on perceptual and cognitive priors should each be consistent across paradigms.

To achieve this, the current study employed six predictive processing tasks measuring healthy participants’ reliance on priors. We first confirmed that the expected prior effects occurred in each task, before submitting these measures of prior strength to a factor analysis. We then planned to relate these latent constructs to psychosis-like experiences by correlating the factor scores with schizotypy metrics. Identifying common factors that underpin predictive processing across diverse paradigms, such as those relating to the hierarchical origin of the prior, and how these relate to subclinical experiences could clarify the mechanisms underlying frank illness and inform the design of tasks for diagnosis and prognosis.

## 2 Methods

### 2.1 Participants

Participants (*N* = 73; 47 female) were recruited using online advertisements and a university volunteers register. Informed consent was obtained at the start of the study. Participants received £30 compensation. All participants were native English speakers aged 18 or above with no history of schizophrenia or psychosis, learning impairment, neurological disorder, or hearing impairment. All participants had normal or corrected-to-normal vision.

### 2.2 Study Procedure

The study was approved by the Cambridge Psychology Research Ethics Committee (PRE.2019.002) and consisted of a pre-experiment online questionnaire followed by two (approximately 2-hr) in-person sessions that, for the majority, took place on separate days (Figure 1). The task order was pseudorandomised: the same tasks always featured in each session, but the order changed to ensure between-task differences were not due to fatigue. The tasks were completed in a quiet room, using noise-cancelling headphones for auditory tasks.

**Figure 1.**
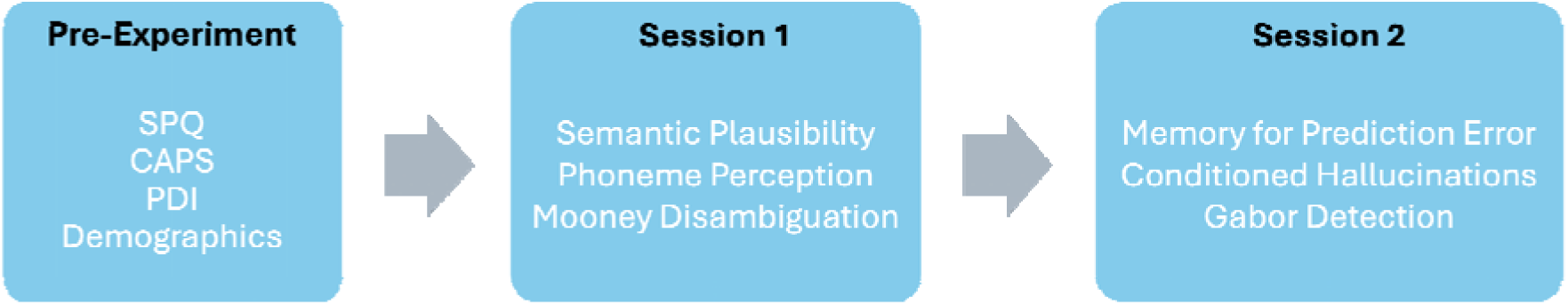
Flow Diagram Illustrating the Study Procedure. The pre-experiment phase consisted of a series of online questionnaires. Sessions 1 and 2 were conducted in-person and involved the completion of six predictive processing tasks. SPQ – Schizotypal Personality Questionnaire^20^; CAPS – Cardiff Anomalous Perceptions Scale^21^; PDI – Peters et al. Delusion Inventory (21-item version).^22^ Alt text: Flow Diagram illustrating the study procedure, listing the pre-experiment questionnaires and the computerised tasks completed in sessions 1 and 2.

### 2.3 Questionnaires

A total score was computed for the schizotypal personality questionnaire^20^ (SPQ), Cardiff Anomalous Perceptions Scale^21^ (CAPS), and Peters et al. Delusion Inventory^22^ (PDI). The CAPS and PDI measure anomalous perceptions and delusions, respectively. Each item requires a (yes/no) report of symptom presence followed by three additional scales (distress, preoccupation/intrusiveness, and conviction/frequency) for endorsed items. As these three scales were highly correlated, we computed severity scores by averaging the sum of the three scales for endorsed items. A summary of schizotypy scores and standardised score distributions are presented in Table 1 and Figure S4, respectively.

**Table 1.** Summary of Demographics and Schizotypy. Severity scores for the CAPS and PDI represent the average sum of the three subscales (distress, preoccupation/intrusiveness, and conviction/frequency) for endorsed items. The three SPQ factor scores are based on Raine et al.^23^. PDI – Peters et al. Delusions Inventory (21-item version)^22^; CAPS – Cardiff Anomalous Perceptions Scale^21^; SPQ – Schizotypal Personality Questionnaire.^20^ Alt text: Table summarising the means and standard deviations for age, SPQ, CAPS, and PDI.

|  | Mean (SD) |
| --- | --- |
| **Age (years)** | 30.56 (13.68) |
| **SPQ** |  |
| Total Score | 18.21 (12.39) |
| Perceptual-Cognitive | 6.25 (5.42) |
| Disorganised | 4.78 (3.82) |
| Interpersonal | 8.97 (6.6) |
| **CAPS** |  |
| Total Score | 3.47 (3.87) |
| Severity | 4.65 (3.54) |
| **PDI** |  |
| Total Score | 3.15 (2.86) |
| Severity | 5.72 (3.67) |

### 2.4 Predictive Processing Tasks

As conflicting results on predictive processing in psychosis originate from diverse paradigms, we chose six tasks from language, visual, memory, and auditory domains. Tasks were selected so that perceptual and cognitive priors featured in the battery. Perceptual priors were defined as inducing implicit and/or automatic lower-level constraints on sensory processing. Cognitive priors induced higher-level beliefs that provided contextual abstractions, involving rapidly learned schemas, semantics, or explicit probabilities. For all tasks, there was existing evidence that the outcomes reflected reliance on priors, and a number of these outcomes had previously been linked to psychosis (see supplementary). Each task produced at least one measure of reliance on priors (Table 2). Full details of each task are presented in the supplementary.

**Table 2.** Predictive Processing Task Battery. Participants completed six predictive processing tasks. The domain, prior type, description of the prior, and parameterisation of the prior effect are detailed for each task. RT – Reaction Time; PE – Prediction Error. Alt text: Table listing details of the domain, prior manipulation, prior type, and parameterisation of the prior effect for each task.

| Task | Domain | Prior Type | Prior | Description of Prior Effect | Parameterisation |
| --- | --- | --- | --- | --- | --- |
| Semantic Plausibility Judgement | Language | Cognitive | Sentence fragment | High-constraint fragments lead participants to expect a specific terminal word. Low-constraint fragments induce broader expectations. | High-constraint prior speeds RT for expected (high-cloze) completions and slows RT for unexpected (low-cloze) completions compared to low-constraint prior. Prior effect indexed by difference in RT between (high- and low-constraint) priors for each sentence completion type. |
| Phoneme Perception |  |  |  |  |  |
| Perceptual Task | Auditory Processing | Perceptual | Videos of articulatory movements | Phoneme perception is biased towards the phoneme indicated by the visual prior. | Change in phoneme category boundary (indifference point) between prior and reference (no prior) conditions |
| Cognitive Task | Auditory Processing | Cognitive | Grapheme–sound associations presented in | Phoneme perception is biased towards the phoneme indicated by the | Change in phoneme category boundary (indifference point) between prior and reference (no |

**Table 2. Predictive Processing Task Battery.** Participants completed six predictive processing tasks. The domain, prior type, description of the prior, and parameterisation of the prior effect are detailed for each task. RT – Reaction Time; PE – Prediction Error.
|  |  |  | training | visual prior. | prior) conditions |
| --- | --- | --- | --- | --- | --- |
| Memory for Prediction Error | Memory | Cognitive | Learned scene–word associations | Effect of PE on memory for scene–word pairs. High PE stimuli, which violate expectations, are associated with better memory recall than (low PE) stimuli that match expectations. | Difference in memory recall between high and low PE conditions |
| Conditioned Hallucinations | Auditory Processing | Perceptual | Learned checkerboard–tone associations | Prior expectations that the checkerboard and tone stimuli are deterministically associated induces perceptions of tones in silence (i.e., conditioned hallucinations) or at subthreshold intensity. | The proportion of ‘tone present’ responses in conditioned hallucination or subthreshold tone conditions. |
| Mooney Disambiguation | Visual Processing | Perceptual | Coloured template image on which the Mooney figure is based | Facilitatory effect of template on disambiguating ambiguous (Mooney) images ($d'$) | Difference in $d'$ between pre- and post-template phases |
| Gabor Detection | Visual<br>Processing | Cognitive | Written<br>information<br>indicating that<br>Gabor presence is<br>likely or unlikely | Effect of prior on response<br>bias for reporting Gabor<br>presence | Difference in response bias<br>between prior and reference<br>conditions |

The *semantic plausibility (judgment) task* measured the effect of sentence constraint, a cognitive prior, on reaction time (RT) for judging the plausibility of sentence completions. High-constraint sentence fragments offer a precise prior, leading participants to expect a specific terminal word. Greater reliance on this prior therefore speeds RTs for the predicted (*high-cloze*) word and also slows RTs for plausible but unexpected (*low-cloze*) words. This task therefore produced two measures of participants’ reliance on priors, comparing the effect of (high versus low) sentence constraint on the RT for judging the semantic plausibility of high-and low-cloze completions.

The *phoneme detection task*^19^ employed separate subtasks to estimate the shift in the /ba/–/da/ category boundary caused by perceptual and cognitive priors. For example, presenting silent videos of an actor articulating /ba/ (perceptual prior) or the written cue ‘BA’ (cognitive prior) increases the proportion of /da/ required in the auditory stimulus to reach the category boundary (known as the *indifference point*) when compared to a reference (‘no prior’) condition. In this task, the prior effect was therefore indexed by the shift in the indifference point when a prior was presented compared to the reference condition.

The *memory for prediction error task*^24^ measured the benefit of surprisal on memory. Participants learned scene–word category associations (a cognitive prior) that were later violated (high PE condition) or maintained (low PE condition). Increased reliance on the prior increases the PE magnitude, enhancing memory performance on high PE trials. Reliance on priors was therefore indexed by the difference in performance between PE conditions.

The *conditioned hallucinations task*^11^ implemented Pavlovian conditioning by pairing a checkerboard with a tone during training. This phase induced a perceptual prior: the belief that the two stimuli were deterministically associated. The effect of this prior was indexed by the likelihood of perceiving tones in silence (i.e., conditioned hallucinations) or at subthreshold intensity, producing two measures for this task.

The *Mooney disambiguation task* measured the improvement in figure-background discrimination (*d*’) of ambiguous images following exposure to unambiguous templates (perceptual prior). Finally, the *Gabor detection task* quantified the effect of written information (cognitive prior) communicating the probability of stimulus presence on response bias. The prior either indicated that the Gabor stimulus was likely to be absent or present, while the reference condition informed participants that both events were equally likely. The prior effect was indexed by the change in response bias for each prior (Gabor present/absent) condition compared to the reference condition, creating two measures for this task.

### 2.5 Statistical Modelling

Analyses were conducted in RStudio^25^ (v4.2.2) and are publicly available (osf.io/9htkf). Initial analyses sought to verify that priors induced the expected effects in each task (see supplementary). The magnitude of the prior effect in each task was then ascertained for each participant. Separate multiple linear regressions were conducted to assess whether all 10 outcomes (measuring reliance on priors) could together predict schizotypy. Separate models were computed for total SPQ, total PDI, total CAPS, PDI severity, and CAPS severity scores and each featured the 10 prior effect outcomes as predictors.

### 2.6 Factor Analysis Model

#### 2.6.1 Assumptions

The 10 outcomes assessing reliance on priors were submitted to an exploratory factor analysis in lavaan.^26^ Pairwise deletion was used in order to maximise the data included in the analysis. The identification of factors requires correlations between clusters of outcomes. To test the suitability of the data for factor analysis, we computed a correlation matrix featuring all outcomes. Spearman’s correlations were used as these assess whether participants ranking high on one measure have similar rankings on other measures instead of reporting linear relationships between outcomes. Unadjusted *p*-values were reported for these correlations as correction factors tend to be overly conservative and limit statistical power. This is particularly problematic in factor analysis designs, which feature multiple outcomes. This was acceptable as the factor analysis was exploratory, aiming to detect possible relationships between outcomes rather than confirming pre-determined hypotheses. Bartlett’s test of sphericity was used to assess whether the correlation matrix significantly differed from an identity matrix. The Kaiser-Meyer-Olkin (KMO) test rated the suitability of the data for factor analysis by measuring sampling adequacy. A reliable factor structure depends on clusters of outcomes that have high loadings on their respective factor(s) and exhibit low cross-loadings (onto other factors). Ideal KMO statistics are closer to 1, while smaller values indicate the existence of partial correlations, which are symptomatic of diffuse correlations.

#### 2.6.2 Factor Extraction

Theoretically, the maximum possible number of factors is equal to the number of outcomes. Factor extraction determines the number of factors that best describes the data. Eigenvalues represent the amount of variance across all outcomes that can be explained by a given factor; for example, an eigenvalue of two indicates that the factor explains the same amount of variance as two task outcomes. We generated a scree plot, which displays the eigenvalues for all possible factors (i.e., for each outcome) in order of magnitude. Large studies (*N* > 200) often use the point of inflexion on the plot as the threshold for the inclusion of factors. Given our relatively small sample, we opted to interpret the plot using Kaiser’s criterion, which recommends that only factors with eigenvalues greater than 1 should be extracted. We validated the scree plot results using a parallel analysis, which is regarded as the optimal method for factor extraction. This method only retains factors with eigenvalues greater than those in a random identity matrix.

#### 2.6.3 Factor Rotation

Factor loadings were estimated using the maximum likelihood method. This was followed by oblimin factor rotation. The sum of squared loadings (i.e., eigenvalue) for each factor was used to interpret whether each factor explained a significant amount of variance.

## 3 Results

### 3.1 Verifying the Effect of the Prior In Each Task

#### 3.1.1 Semantic Plausibility Task

Results for the semantic plausibility task revealed the expected effects of sentence constraint on RT for judging the semantic plausibility of the terminal words (Figure 2C). Mean RTs for high-cloze (predicted) completions were significantly faster in high-(*M* = 0.63, *SD* = 0.09) versus low-constraint (*M* = 0.69, *SD* = 0.07) contexts (*b* =-0.06, *SE* = 0.02, *z* =-3.78, *p* <.001; *d* = 0.19). This *high-cloze prior effect* represents the benefit of a precise prior for judging the plausibility of the most likely sentence completion. In contrast, slower RTs were observed for low-cloze completions in high-(*M* = 0.96, *SD* = 0.12) versus low-constraint (*M* = 0.85, *SD* = 0.09) contexts (*b* = 0.11, *SE* = 0.04, *z* = 2.95, *p* =.003; *d* = 0.33). This *low-cloze prior effect* reflects the detrimental effect of a precise prior on judging plausible but unexpected sentence completions.

**Figure 2.**
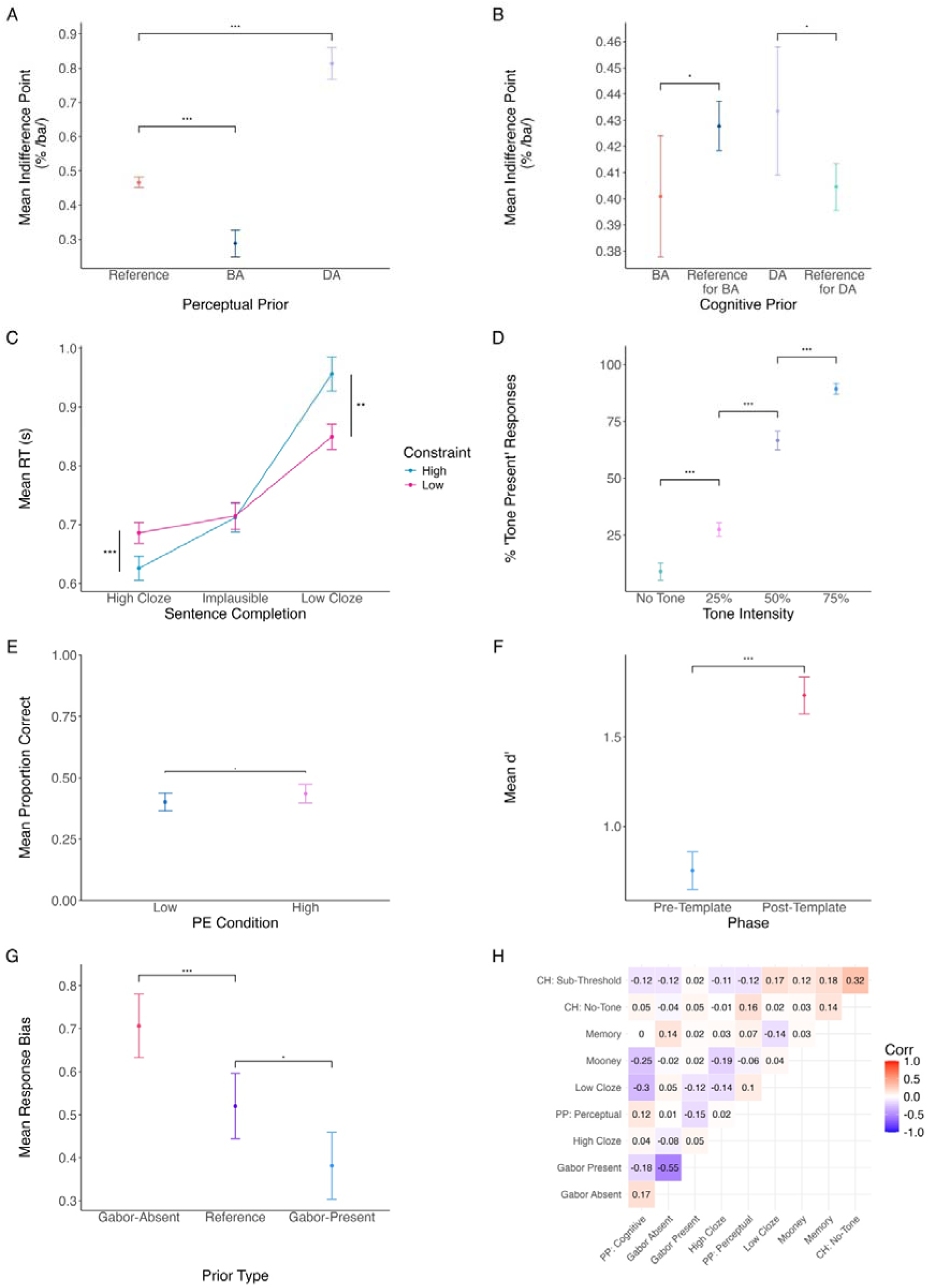
Prior Effects in the Predictive Processing Tasks. A–B) Phoneme Perception (PP): Mean indifference points for the BA and DA conditions each significantly differed from the reference condition in both perceptual and cognitive subtasks. C) Semantic Plausibility Judgment: High-constraint contexts speeded RTs for high-cloze completions and slowed RTs for low-cloze completions. D) Conditioned Hallucinations (CH): The percentage of tones endorsed increased with tone intensity. E) Memory for PE: There was a borderline-significant improvement in memory for stimuli on high PE trials. F) Mooney Disambiguation: Discrimination performance (*d*’) was significantly enhanced in the post-template phase. G) Gabor Detection: Priors indicating Gabor absence decreased the response bias criterion, while priors indicating Gabor presence liberalised this criterion. H) Spearman’s correlation matrix for the 10 measures of reliance on priors. All error bars represent 95% CIs, adjusted to account for within-participant variance.^27^ PE – Prediction Error; RT – Reaction Time. •*p* <.01, \**p* <.05, \*\**p* <.01; \*\*\**p* <.001 Alt text: Grid of figures illustrating the prior effect in each task. The final figure displays a correlation matrix of task scores.

#### 3.1.2 Phoneme Perception Task

Results from the phoneme perception task revealed the expected effects of both the perceptual and cognitive priors on the indifference point (Figures 2A–B), which was defined by the weighting of /ba/ (ω_BA_) in the auditory stimulus. In the perceptual task, articulatory movements for /ba/ (BA condition) reduced the average indifference point (*M* =.29, *SD* =.18) compared to the reference condition (*M* =.47, *SD* =.12; *b* =-0.18, *SE* = 0.03, *t* =-6.97, *p* <.001; *d* = 1.17). In contrast, the average indifference point for the DA condition (*M* =.81, *SD* =.21) was significantly greater than the reference (*b* = 0.35, *SE* = 0.03, *t* = 13.59, *p* <.001; *d* = 3.45). The cognitive task priors produced similar patterns of results. The ‘BA’ graphemes (BA condition) significantly reduced the indifference point (*M* =.40, *SD* =.12) compared to the corresponding (BA) reference condition (*M* =.43, *SD* =.15; *b* =-0.03, *SE* = 0.01, *t* =-2.10, *p* =.037; *d* = 0.35). The DA graphemes (DA condition) significantly increased the indifference point (*M* =.43, *SD* =.11) when compared with the corresponding (DA) reference condition (*M* =.40, *SD* =.11; *b* = 0.03, *SE* = 0.01, *t* = 2.27, *p* =.024; *d* = 0.38). The magnitude of the shift in indifference point for the DA and BA priors were summed to produce a single metric of prior strength for each subtask to be submitted for factor analysis.

#### 3.1.3 Conditioned Hallucinations Task

As expected, tones presented at higher intensities were endorsed more frequently in the conditioned hallucinations task (Figure 2D). 75% threshold trials featured more tone reports (*M* =.89, *SD* =.09) than 50% trials (*b* = 1.57, *SE* = 0.12, *z* = 13.10, *p* <.001; *OR* = 0.21, *M* =.67, *SD* =.15), which in turn featured more tone reports than 25% trials (*M* =.27, *SD* =.11; *b* = 2.01, *SE* = 0.10, *z* = 19.45, *p* <.001; *OR* = 0.13). Tone reports were significantly less likely in no-tone (*M* =.09, *SD* =.13) versus 25% trials (*b* = 1.68, *SE* = 0.16, *z* = 10.54, *p* <.001; *OR* = 0.19). The percentage of tone endorsements in the no-tone (i.e., conditioned hallucinations) and subthreshold trials represented the effect of priors and were entered into the factor analysis as separate effects.

#### 3.1.4 Memory for Prediction Error Task

Analyses of the initial three phases of the memory for PE task demonstrated that participants successfully learnt the scene–word category associations (priors): Significant intercepts (all *p*s <.001) indicated above-chance accuracy in familiarisation, training, and study phases. In testing, mean memory performance across participants (*M* =.42, *SD* =.11) was above chance (.33). Surprisingly, there was no significant effect of PE condition (*p* =.079; Figure 2E). Excluding self-reported guesses did not alter this result. Given the similarity of the effect size to Greve et al.’s^24^ result, this finding could be due to a lack of power. An identical model with all testing trials included (i.e., not excluding trials with stimuli that were incorrectly predicted in the study phase; see supplementary) produced a significant effect of PE condition (*b* = 0.17, *SE* = 0.09, *z* = 2.00, *p* =.046), with superior memory for high-(*M* =.44, *SD* =.13) versus low (*M* =.40, *SD* =.14) PE trials. There was a similar difference in performance between high-(*M* =.43, *SD* =.13) and low (*M* =.40, *SD* =.15) PE trials in the original model (with incorrectly predicted trials excluded). Furthermore, an analysis of the incorrectly predicted trials did not show a significant effect of PE condition on memory, hence the condition effect was not driven by these trials. Overall, this suggests superior memory for high PE stimuli, but a lack of power to detect this effect.

#### 3.1.5 Mooney Disambiguation task

The Mooney disambiguation task produced the expected (prior) effect of template exposure on *d*’, with higher d’ in the post-template (*M* = 1.73, *SD* = 0.86) than pre-template (*M* = 0.75, *SD* = 0.44) phase (Figure 2F; *b* = 0.98, *SE* = 0.07, *t* = 13.26, *p* = <.001; *d* = 2.29).

#### 3.1.6 Gabor Detection Task

In the Gabor detection task, priors communicating the probability of stimulus presence successfully influenced response bias (Figure 2G). The prior suggesting Gabor absence produced a more conservative criterion (*M* = 0.71, *SD* = 0.53) compared to the reference condition (*M* = 0.52, *SD* = 0.45; *b* = 0.19, *SE* = 0.05, *t* = 3.47, *p* <.001; *d* = 0.61). The prior suggesting Gabor presence produced a more liberal criterion (*M* = 0.38, *SD* = 0.49) compared to the reference condition (*b* = 0.14, *SE* = 0.05, *t* = 2.58, *p* =.011; *d* = 0.46).

### 3.2 Factor Analysis

The dataset featured 73 participants, of which 40 had complete data on all task measures. The use of Kaiser’s criterion as a threshold indicated extraction of a single-factor structure, while the parallel analysis recommended a two-factor structure. We computed the factor analysis with both models and compared the amount of variance explained by each. The factor analysis reported eigenvalues greater than 1 for the two factors, indicating that the two-factor model explained significantly more variance (26%) than the single-factor model (14%).

The two-factor model loadings are presented in Table 3. As standard, factor loadings greater than 0.3 are deemed significant. The first factor had the highest loadings for the effects of the Gabor-present prior and Gabor-absent prior on response bias. The second factor had the highest loading for the measure of tone reports on subthreshold trials in the conditioned hallucinations task. The measure of conditioned hallucinations (i.e., reporting tone presence on tone-absent trials) also loaded onto this factor with borderline significance. Both factors therefore represent within-task correlations rather than a consistent weighting of priors across different tasks.

**Table 3.** Factor Loadings for the Two-Factor Model of Reliance on Priors. Significant loadings (> 0.3) are shown in **bold**. Factor loadings in this model are regression coefficients. Alt text: Table demonstrating the results of the factor analysis: a two-factor model.

|  | Prior Type | Factor 1 | Factor 2 |
| --- | --- | --- | --- |
| **Conditioned Hallucinations** |  |  |  |
| Subthreshold Trials | Perceptual | 0.19 | **0.97** |
| Tone-Absent Trials | Perceptual | 0.12 | 0.29 |
| Memory for Prediction Error | Cognitive | 0.06 | 0.28 |
| **Semantic Plausibility** |  |  |  |
| High-Cloze | Cognitive | 0.14 | 0.03 |
| Low-Cloze | Cognitive | -0.10 | 0.25 |
| Mooney Disambiguation | Perceptual | -0.03 | 0.11 |
| **Gabor Detection** |  |  |  |
| Gabor-Absent Prior | Cognitive | **-0.52** | 0.04 |
| Gabor-Present Prior | Cognitive | **0.98** | -0.26 |
| **Phoneme Perception** |  |  |  |
| Perceptual Prior | Perceptual | -0.20 | -0.02 |
| Cognitive Prior | Cognitive | -0.16 | -0.02 |

Overall, tests of assumptions indicated that the data were unsuited to factor analysis, and therefore the two-factor model may be unreliable. First, few outcomes were significantly correlated (Figure 2H), despite the use of Spearman’s correlations, which tend to be larger as they measure whether relationships between variables are monotonic. This was reflected in Bartlett’s test (*p* =.07), which indicated that the correlation matrix did not significantly differ from an identity matrix. Second, the KMO test revealed that the overall measure of sampling adequacy was ‘unacceptable’ (.44). According to standard methods, such results may be corrected by the removal of outcomes with statistics less than.5; however, only two variables exceeded this threshold. This indicated diffuse correlations rather than clusters of correlations representing factors. Although these violations limit the reliability of the two-factor model, importantly they highlight how individuals may not implement a consistent strategy for weighting priors across tasks (see discussion).

### 3.3 Modelling Schizotypy as a Function of Prior Effects

As the factor analysis did not produce reliable factors representing reliance on priors across tasks, we were unable to conduct the planned correlations between schizotypy and factor scores. Instead, we investigated whether performance across the task battery could successfully predict schizotypy. We modelled schizotypy as a function of *all of the measures* of reliance on priors (see supplementary). This model explained significantly more variance (53.6%) than an intercept-only model in the case of PDI severity, *F*(10,29)= 3.35, *p* =.005, but not in the case of other schizotypy measures (full results in Tables S2–6). Higher PDI severity was associated with weaker priors in the phoneme perception (perceptual) and memory for PE tasks. PDI severity was also positively associated with an increased proportion of conditioned hallucinations and a greater effect of semantic constraint for judging the plausibility of low-cloze sentence completions, both indicating an increased reliance on priors.

## 4 Discussion

Dysfunctional predictive processing is a frequently cited theory of psychosis. Authors tend to generalise findings from a single task to advance a theory of either strong or weak priors in psychosis. However, there is no evidence that the weighting of priors across tasks is underpinned by a single factor, and correlations with psychosis often produce conflicting results. While some research has suggested that a hierarchical model of (perceptual vs cognitive) priors may reconcile findings,^18,19^ this has not been evaluated across paradigms until now. The present study conducted a factor analysis to assess whether this two-factor model underpinned performance on six predictive processing tasks and evaluated whether performance across tasks predicted schizotypy.

Although a two-factor model best described performance across tasks, these factors represented prior strength in the conditioned hallucinations and Gabor detection tasks, respectively; outcomes from other tasks did not significantly load onto either factor. As such, the model only accounted for 26% of variance in performance and indicated that the weighting of priors was task-specific. Furthermore, even within-task measures did not always correlate as expected, limiting the interpretability of the results. In the Gabor detection task, measures of reliance on the stimulus-present and stimulus-absent priors were negatively correlated, indicating that participants do not necessarily show equivocal reliance on different priors even within a task. This task exemplifies how the content of contextual information can determine the weighting of the prior. Participants displayed a response bias to report stimulus absence across conditions, therefore priors aligning with this ‘baseline’ belief may have been more readily incorporated into decision-making. Meanwhile, as expected, the prior exerted different effects according to the type of stimulus in the semantic plausibility task (high-vs low-cloze words), yet surprisingly the magnitude of these effects were not correlated. In other words, a greater effect of semantic constraint on performance with high-cloze completions was not indicative of a greater effect with low-cloze completions. Hence, the weighting of priors can depend on the specific stimuli.

Our results serve to caution authors against extrapolating results from a single task across paradigms to advance a general theory of dysfunctional predictive processing in psychosis. The two-factor model was associated with high partial correlations and weakly correlated task outcomes, indicating limited shared variance. While small samples reduce the precision of correlation estimates, they do not preclude their detection^28^; thus, the lack of meaningful intercorrelations is unlikely to be explained solely by sample size. Instead, our results suggest that individuals may not implement a consistent strategy for weighting priors across tasks. These factor analysis results are based on this specific dataset and may not replicate with different tasks. A previous^29^ factor analysis of predictive processing that focused solely on the visual domain identified a two-factor structure that explained significantly more variance (50%) than our model, although this dataset also violated assumptions. Our battery tested predictive processing across domains, and therefore the lack of a reliable factor structure and poor explanatory power could partly reflect the diversity of tasks. Future research could evaluate whether domain-specific hierarchical models can explain variations in performance. For example, in the visual domain, Lhotka et al.^30^ reported that distinct (bistable and pareidolia) illusions linked to predictive processing are underpinned by separate factors. Specifically, pareidolia was governed by higher-level cognitive processes than bistable illusions. It is also important to consider alternative hierarchical models that may present with different task batteries; for instance, the distinction between contextual priors, which operate in isolated spatio-temporal situations, and structural priors, which constrain multiple aspects of perception.^31^

As no reliable factors underpinned performance across tasks, we were unable to correlate factor scores with schizotypy. Instead, we evaluated whether all measures of prior strength from the task battery could predict schizotypy. This only produced a significant model for PDI severity, which was driven by a specific subset of task scores. Higher PDI severity was associated with weaker perceptual priors in the phoneme perception paradigm. The original study with this task^19^ also reported weaker perceptual priors relating to PDI scores in first-episode psychosis. Other studies have shown weaker lower-level priors linked to subclinical^18^ and psychotic^32^ delusional conviction. However, we identified that PDI severity was also associated with an increased susceptibility to conditioned hallucinations, evidencing an increased reliance on (perceptual) priors. An increased susceptibility for conditioned hallucinations has previously been linked to auditory-verbal hallucinations rather than delusional severity.^11,33^ Indeed, (supplementary) regressions indicated 1) a borderline-significant positive association between reports of everyday auditory-verbal hallucinations and conditioned hallucinations and 2) a significant positive association between total CAPS and the likelihood of detecting subthreshold intensity tones. However, after controlling for prior effect outcomes from all other tasks, neither outcome from the conditioned hallucinations task significantly predicted CAPS scores. PDI severity was also related to a decreased reliance on priors in the memory for PE task. This task was not previously implemented in psychosis research, and the reason for this finding is unclear. Finally, higher PDI severity was associated with a greater prior effect on judging the plausibility of low-cloze sentence completions specifically. A similar study^14^ also reported stronger priors associated with SPQ scores, but only under conditions of uncertainty.

Our findings defy previous assumptions that outcomes from different paradigms estimate a unitary metric of prior strength. Multiple studies have generalised results from one task to advance a theory of globally strong or weak priors in psychosis. Generalisations of results assume that individuals’ susceptibility to predictive cues are consistent across diverse tasks, yet in the present data this has no evidential basis. Not only does the relationship between psychosis and predictive processing vary across tasks, but different tasks do not always appear to measure the same ‘prior-weighting’ construct. The comparability of estimates across tasks is limited as different tasks operationalise the weighting of priors using different metrics. In future, computational models may be useful for implementing a common parameter estimation method across tasks. Such models could also track the trial-by-trial updating of priors at different hierarchical levels. This is preferential to the use of aggregate metrics that could mask adaptations in the weighting of priors that develop from experience with the paradigm. Another challenge for this field concerns the design of tasks that isolate predictive processing mechanisms. Although we selected paradigms that had been designed to measure reliance on priors, alternative explanations for results exist. For instance, individual differences in performance in the phoneme detection task could reflect audiovisual integration ability or the prioritisation of auditory or visual input. A previous study^34^ of the McGurk effect reported that psychosis patients display a preference for auditory stimuli, which in this paradigm would be misinterpreted as a reduced reliance on (visual) priors. Future research should also manipulate sensory precision to evaluate whether this modulates reliance on priors. For example, previous research has reported that psychosis-associated changes in the weighting of priors are only apparent under conditions of uncertainty induced by sensory degradation or varying the precision of priors.^14,35^

Overall, this study’s factor analysis does not provide evidence that an individual’s performance across predictive processing tasks can be described by the weighting of priors at two distinct hierarchical levels. As such, inconsistent findings on the relationship between psychosis and predictive processing cannot alone be reconciled by this framework. Further investigation is required to confirm whether additional variables, such as task modality, can improve the predictive validity of this model. This can be achieved by further work implementing multi-paradigm approaches rather than single tasks, combined with computational estimates of reliance on priors that are comparable across paradigms.

## Author Contributions

C Miller-Silva: Conceptualisation, data curation, formal analysis, funding acquisition, investigation, methodology, project administration, software, supervision, visualisation, writing – original draft

F Knolle: Conceptualisation, formal analysis, methodology, software, supervision, writing – review & editing

A Greve: Formal analysis, methodology, resources, software

F de Beer: Data curation, formal analysis, investigation, methodology, writing – review & editing

T Mujirishvili: Data curation, formal analysis, investigation, methodology LJ MacGregor: Conceptualisation, methodology, writing – review & editing PR Corlett: Formal analysis, methodology

J Haarsma: Formal analysis, software

AR Powers: Formal analysis methodology

GK Murray: Conceptualisation, funding acquisition, methodology, project administration, supervision, writing – review & editing.

## Supporting information

Supplementary Methods & Results

## Data Availability

All data produced in the present study are available upon reasonable request to the authors.

