## Supplementary Methods & Results for "Factor Analysing Predictive Processing: No Evidence for a General Factor Across Tasks"

### 1 Over-Arching Statistical Methods

The significance threshold (alpha) was set as 5% unless otherwise stated. For repeated measures designs, mixed-effects modelling was implemented via the lme4 package^1^ (v1.1.34) to avoid pseudoreplication given that the data were non-independent (see Jaeger^2^ for further discussion). All mixed models were optimised using the Bound Optimization BY Quadratic Approximation^3^ (BOBYQA). While maximal models (i.e., with a fully specified random-effects structure) are advised at least for confirmatory hypothesis testing,^4,5^ often these models fail to converge or warn of singularity (overfitting). Hence, the buildmer package^6^ (v2.9) was employed to construct well-specified yet parsimonious models by identifying the largest random-effects structure that produced convergence. Categorical and continuous predictors were grand-mean centered for analyses involving interaction effects in order to reduce (multi-) collinearity between main effects and interactions.

Supplementary analyses investigated the relationship between each task outcome (i.e., measure of reliance on priors) and schizotypy. Five separate regression models were estimated for each task outcome, regressing the effect on each schizotypy predictor individually (total SPQ, total PDI, total CAPS, PDI severity, and CAPS severity).

One participant was excluded from analyses involving schizotypy measures as they did not complete the questionnaire.

### 2 Semantic Plausibility Judgment Task

#### 2.1 Background

The semantic plausibility (judgment) task assessed the use of priors in lexical prediction during reading comprehension. The initial part of a sentence induces expectations about the terminal word, especially if this sentence fragment offers a highly constrained semantic context (e.g., ‘The boy went to the beach with his bucket and *spade*’). Constrained context (i.e., a precise prior) speeds the processing of predicted words (that match prior expectations) but slows processing of plausible yet unexpected items.^7^ In a recent study,^8^ participants heard sentences with different degrees of semantic constraint and reported the final word, which was distorted by noise. Results revealed that schizotypy was associated with a greater reliance on semantic expectations.

#### 2.2 Paradigm

The semantic plausibility task measured participants’ reliance on lexical predictions when judging the plausibility of sentence completions. The task was programmed in MATLAB R2018b using the Psychophysics Toolbox Version 3^9^ and consisted of 5-min training and 25-min testing phases. In both phases, participants were presented with written sentences appearing onscreen one word at a time and produced speeded judgments of the semantic plausibility of the terminal word. Words appeared for 300 ms, with each word replacing the previous word. The final word was succeeded by a (red dot) response cue. Participants reported whether the terminal word was plausible (right arrow key) or implausible (left arrow key) given the sentence context. Response keys were labelled in order to reduce illegal key presses.

The sentence stimuli were taken from Federmeier et al.^10^ The *cloze probability* (predictability of a word in a given context) of the terminal word varied according to six task conditions (Table S1), which were defined by the semantic *constraint* of the sentence and the plausibility of the final word. *Implausible* completions did not feature in the original stimuli and were therefore generated and independently checked for plausibility and grammaticality by three researchers. *High-constraint* sentences led participants to expect a specific terminal word, while *low-constraint* sentences induced broader predictions. The initial sentence fragment therefore induced prior beliefs about the identity of the terminal word. High- and low-constraint sentences were matched in length.

This task generated two prior effect measures: the effect of constraint on reaction time (RT) for assessing the plausibility of 1) *high-cloze* (most predictable) and 2) *low-cloze* (plausible but unexpected) completions. Specifically, we hypothesised that higher constraint would speed RTs for high-cloze completions and slow RTs for low-cloze completions.

The training phase consisted of 10 trials and was designed to familiarise participants with the task. This phase featured three sentences that were repeated to present an equal number of high- and low-constraint trials and sentence completions from all three conditions. The testing phase consisted of 240 trials and featured two breaks to reduce fatigue. This phase used 80 sentences (with equal proportions of high- and low-constraint items), each presented three times (with high-cloze, low-cloze, and implausible completions). The order of stimuli was randomised for constraint, plausibility, and sentence item.

| Constraint | Sentence  Completion | Example | Mean Cloze  Probability |
| --- | --- | --- | --- |
| High Constraint |  | Father carved the turkey with a ... |  |
|  | High cloze | knife | .85 |
|  | Low cloze | smile | .009 |
|  | Implausible | plant | 0 |
| Low Constraint |  | In the wooden box the girl found a ... |  |
|  | High cloze | ring | .29 |
|  | Low cloze | doll | .015 |
|  | Implausible | pressure | 0 |

**Table S1. Examples of the Six Semantic Plausibility Task Conditions.** High-cloze completions in high-constraint sentences were the frequently predicted words for those contexts. High-cloze completions in low-constraint sentences were the most frequently predicted words but had lower cloze probability ratings than in high-constraint contexts. Low-cloze completions were semantically plausible but were less predictable from the sentence context. Implausible completions had a cloze probability of zero.

#### 2.3 Data Pre-Processing

All participants (*N* = 70) performed above chance in training and therefore understood the task. One sentence was identified as an outlier in terms of the percentage of incorrect trials across all participants; this item featured a spelling error, hence all trials featuring this sentence were removed (210 responses; 1.25% of the data).

It was important to eliminate trials that featured implausibly fast RTs (e.g., rushed responses). Previous literature with lexical decision tasks suggests that sub–200-ms RTs should be excluded.^11^ It was expected that these trials would be equally distributed across all six conditions and mostly feature incorrect responses. However, sub–200-ms RTs were more frequent for high-cloze completions in high-constraint contexts and were more likely to be correct, therefore instead the RT threshold was titrated to ensure a 66% (chance) response accuracy on these fast trials. This meant that RTs faster than 70 ms were excluded from the analyses (143 trials; 0.9% of the remaining data). A sub–200-ms threshold was also employed by a study with a similar paradigm.^12^

As responses were speeded, relatively fast RTs were expected. It was important to remove trials that had unusually slow RTs, indicating that participants may have been distracted. Minimal outlier trimming, following the procedure by Haeuser & Kray,^12^ was used to eliminate trials where the RT was three times greater than the interquartile range (> 2.6 s; 481 responses, 3% of the remaining data). This ensured the exclusion of extreme values with minimal data loss.

Incorrect responses were also removed (1,122 responses; 7% of the remaining data). This ensured that the results isolated the effect of the prior on RT without performance accuracy as a confound.

#### 2.4 Exploring the Effect of Constraint on Judging Semantic Plausibility

A linear mixed-effects model investigated the effect of sentence constraint on RT for the three types of sentence completions (high cloze, low cloze, implausible). Simple coding (for completion type) and dummy coding (for constraint) were used to create pre-planned contrasts to analyse how mean RT varied across conditions. This method is preferred to *t*-tests, which present with the problem of multiple comparisons and do not generalise well to mixed models.^13^ However, due to constraints on the number of planned contrasts that can be assigned to a variable in a single analysis, post-hoc *t*-tests were used to explore significant interactions. *P*-values for these analyses were corrected using the Tukey method. Due to non-normal residuals and heteroscedasticity, the bestNormalize package^14^ (v1.9.1) was used to choose the optimal transformation for the data, in this case the Ordered Quantile normalisation. The results report untransformed statistics for ease of interpretation. The random-effects structure included random intercepts for participants, sentences, and trial number; by-participant slopes for completion type and constraint; and by-item slopes for completion type.

#### 2.5 Supplementary Analyses: Reliance on Priors as a Function of Schizotypy

A series of linear regressions investigated the relationship between each effect of the semantic prior, namely high- and low-cloze prior effects, and schizotypy. There was no significant relationship between any of the schizotypy measures and either prior effect.

### 3 Phoneme Perception Task

#### 3.1 Background

The phoneme perception task^15^ employed separate subtasks to measure how perceptual and cognitive visual priors affect phoneme perception. Haarsma et al.^15^ suggested that weakened perceptual priors in the at-risk mental state (ARMS) were compensated by a strengthening of cognitive priors that becomes evident in first-episode psychosis (FEP). There was also a positive correlation between cognitive prior strength and delusions in ARMS and a negative correlation between perceptual prior strength and both delusions and hallucinations in FEP. This may evidence how the observed changes in predictive processing in these groups convert to symptoms: the compensatory increase in cognitive prior strength may induce delusions, and a failure by this mechanism to increase perceptual prior strength may cause symptoms to persist. In contrast, psychosis-like symptoms in controls were unrelated to the task results.

#### 3.2 Paradigm

Full details of the paradigm can be found in Haarsma et al.^15^ Participants judged the identity of ambiguous phonemes, which were a combination of /ba/ and /da/. The weighting of /ba/ (ω_BA_) in the phoneme varied in a psychophysical staircase that converged on the *indifference point*, which represented category boundary, or the stimulus that participants found maximally ambiguous. Visual priors (Figure S1) biased phoneme perception and consisted of videos of articulatory movements for /ba/ or /da/ (perceptual task) or graphemes that had deterministically predicted the phonemes in training (cognitive task). The influence of the priors on perception was assessed by comparing the shift in indifference point for each (BA or DA) prior condition relative to the reference (no prior) condition. For example, we expected that a greater ω_BA_ would be required to overcome the effect of the DA prior (increased indifference point relative
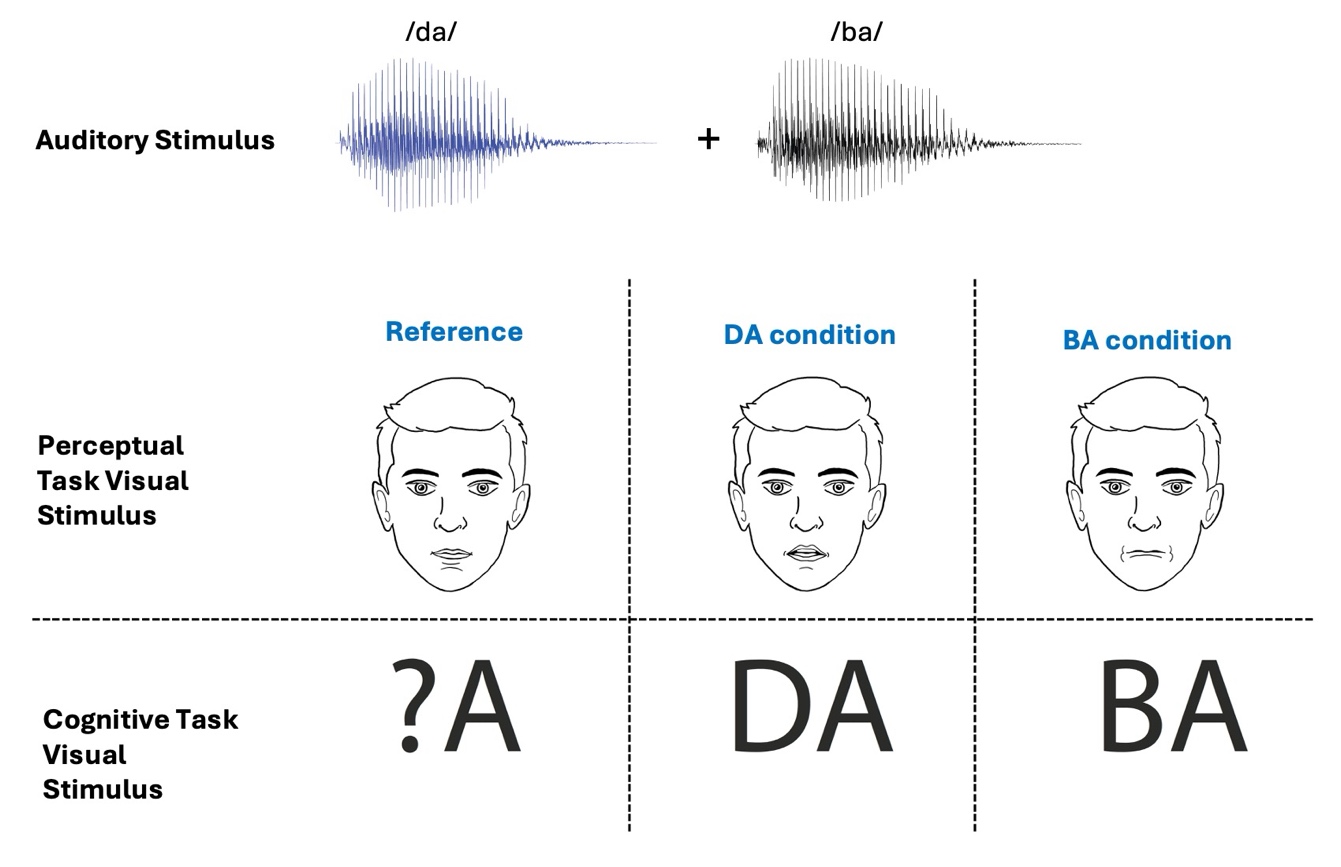
to the reference).

**Figure S1. Illustration of the Phoneme Perception Task Conditions.** Auditory stimuli were a combination of /ba/ and /da/ phonemes. Priors influencing perception were induced by videos of articulatory movements for /ba/ or /da/ (perceptual task) or written cues that had deterministically predicted the phonemes in training. Reference (no prior) conditions had an image of a still face (perceptual task) or a ‘?A’ written cue (cognitive task). Perceptual priors were induced using videos of a real person: the cartoon face shown here is for demonstration purposes only.

#### 3.3 Exploring the Effect of Priors on the Indifference Point

Seventy-one participants completed both tasks. Linear mixed-effects models, with random intercepts for participants, were used to confirm the expected prior-responsive shifts in the indifference point. As the perceptual task featured two staircases (with different initial ω_BA_ values) for each prior condition, Pearson’s correlations were used to confirm that the staircases reliably approximated the same indifference point. These measures were therefore averaged for each prior condition and submitted to the mixed model. For the perceptual task, the (dummy-coded) contrasts compared the indifference point for the reference condition with the BA and DA conditions separately. For the cognitive task, user-defined contrasts^13^ were created to compare mean indifference points for the 1) BA versus BA reference and 2) DA versus DA reference conditions. Sensitivity analyses excluded cases of priors dominating perception (indifference points of one or zero) and terminated staircases (due to time constraints) to confirm that these values, which were not true indifference points, did not significantly distort the results. The total prior effect for each subtask, which was the measure submitted for factor analysis, was the sum of the BA and DA prior condition effects.

#### 3.4 Supplementary Analyses: Reliance on Priors as a Function of Schizotypy

Weaker perceptual priors were associated with increased PDI severity (*b* = -0.02, *SE* = 0.01, *t* = -2.32, *p* = .023) and total SPQ (*b* = -0.01, *SE* = 0.002, *t* = -2.14, *p* = .036). Separate follow-up analyses with each SPQ factor score as a predictor revealed that the latter effect was mainly driven by interpersonal traits (*b* = -0.01, *SE* = 0.01, *t* = -2.67, *p* = .01; Raine et al., 1994). There was no association between cognitive prior strength and schizotypy.

### 4 Conditioned Hallucinations Task

#### 4.1 Background

The conditioned hallucinations task^17^ used proneness to task-induced hallucinations of auditory tones as a measure of reliance on priors. In the original study, computational modelling revealed that susceptibility to these conditioned hallucinations is underpinned by hyper-precise priors in both clinical and non-clinical voice-hearers. This was replicated with a large (*N* = 458) heterogenous cohort of individuals with unusual perceptual experiences, including psychosis-spectrum and non-clinical groups,^18^ which also revealed that susceptibility to conditioned hallucinations correlated with the frequency of auditory hallucinations in daily life.

#### 4.2 Paradigm

Full details of the conditioned hallucinations task can be found in Powers et al.^17^ During training, participants were repeatedly presented with a checkerboard paired with a tone. This induced the belief (prior) that these stimuli are always presented concurrently. In testing, the checkerboard was presented alone (no-tone trials) or alongside tones at different intensities (75%, 50%, or 25% tone thresholds, estimated for each individual using the QUEST procedure). A greater reliance on priors is evidenced by reports of hearing tones on subthreshold (25%) or tone-absent trials.

#### 4.3 Data Pre-Processing

Fifty-six participants completed the task. Four participants formed a distinct group of outliers who missed a substantial number (at least 16%) of trials and were therefore excluded. These participants likely found response timing challenging; indeed, two participants reported difficulties after the experiment. One further participant was excluded as they reporting hearing the tone on only 41% of high-intensity (75%) trials. Thus, 51 participants were submitted to the final analyses.

#### 4.4 Analysing the Effect of Tone Intensity on Reporting Tone Presence

A logistic mixed-effects model was employed to analyse the likelihood of ‘tone present’

reports for trials with different tone intensities. This model included a fixed effect of condition, random intercepts for participants, and by-participant random slopes for condition. Repeated contrasts were used for the condition variable to produce 1) no-tone versus 25%, 2) 25% versus 50%, and 3) 50% versus 75% condition comparisons.

#### 4.5 Supplementary Analyses: Reliance on Priors as a Function of Schizotypy

The relationship between schizotypy and task performance was investigated using a series of logistic mixed-effects models for 25% and no-tone trials separately. These estimated the likelihood of a reporting tones given each schizotypy measure and included random intercepts for participants. There were no relationships between any of the schizotypy scores and the probability of conditioned hallucinations (no-tone trials). However, there was evidence for more subthreshold tone endorsements with higher CAPS severity (*b* = 0.08, *SE* = 0.03, *z* = 2.50, *p* = .012). These results were unaffected by the inclusion of the 75% tone threshold and the number of training trials as covariates in a second-level model.

As the original study^17^ reported that clinical and non-clinical voice-hearers display a greater susceptibility to conditioned hallucinations in this task, we also investigated the relationship between reports of daily auditory-verbal hallucinations and task performance. A binary coding system was used to categorise participants as either ‘voice-hearers’ or ‘controls’. Participants were coded as voice-hearers if they responded in the affirmative to at least one of the following questions from the CAPS:

1. 'Do you ever hear voices commenting on what you are thinking or doing?',

2. 'Do you ever hear voices saying words or sentences when there is no-one around that might account for it?', and

3. 'Have you ever heard two or more unexplained voices talking with each other?’

This categorised 15 participants as voice-hearers. Logistic mixed modelling revealed a borderline significant relationship between voice-hearing in everyday life and conditioned hallucinations (*b* = 0.54, *SE* = 0.28, *z* = 1.94, *p* = .053) only after controlling for the significant negative correlation between conditioned hallucinations and 75% tone threshold.

### 5 Memory for Prediction Error Task

#### 5.1 Background

The memory for prediction error (PE) task was previously used to demonstrate enhanced memory for stimuli that violate expectations.^19^ This result was framed in terms of a predictive processing account,^20^ where unexpected stimuli generate PEs, which in turn drive updates to the generative model (entailing memory encoding). An increased reliance on priors generates larger PEs, which in turn improve memory encoding. This account has, to the best of our knowledge, not until now been assessed in the context of psychosis.

#### 5.2 Paradigm

Full details of the task can be found in Greve et al.^19^ Stimuli were generated using E-Prime 2.0.^21^ We replaced the original (emotion) stimuli with the categories ‘mammals’ and ‘fruit/veg’ to account for possible perturbations in processing valence in schizotypy.^22^ Each category consisted of 28 words that were similar in frequency and length and were independently assessed by two researchers for inclusion in the task. Scene stimuli were photographs, with six distinct images for each of the 48 scenes (288 images in total). Multiple scenes and specific images were changed in line with the new word categories to ensure that no images featured people, animals, fruit, or vegetables to avoid learning biases for the scene–word category pairings.

Across task phases (Figure S2), participants learnt associations between scenes and word categories, which they used to predict upcoming stimuli. In the original study, participants displayed enhanced memory recall for items from high PE trials, where prior beliefs about scene–word category pairings were violated, compared to items from (low PE) trials that conformed to expectations. As a greater reliance on priors would increase the magnitude of the PE, reliance on priors was parameterised as the difference in memory performance for high- versus low PE trials.

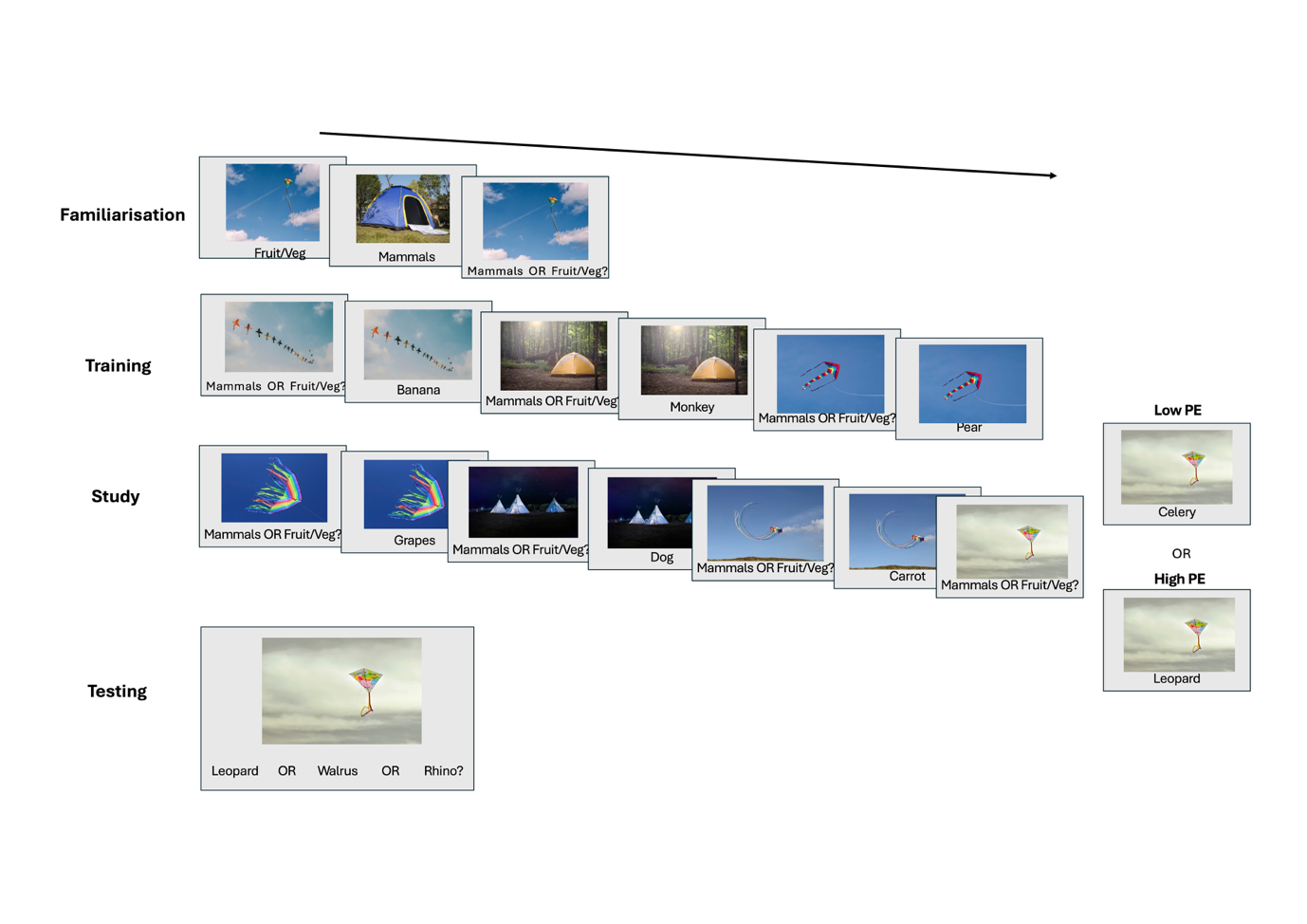

**Figure S2. The Memory for Prediction Error Task.** Familiarisation taught participants scene–word category associations (e.g., kites presented with ‘fruit/veg’). In training, these associations were generalised to novel images from the same scenes and feedback consisted of specific words from the correct category (e.g., a novel scene of kites paired with ‘banana’). The study phase consisted of the same procedure as training with three novel images of each scene, but participants attended to the specific words provided as feedback. The first two images of each scene always conformed to expectations (e.g., kites paired with ‘grapes’ or ‘carrot’). The third image of each scene was used in critical trials, where expectations were either maintained (e.g., kites paired with ‘celery’; low PE condition) or violated (e.g., kites paired with ‘leopard’; high PE condition). The testing phase assessed participants’ memory for image-word pairs presented in the study phase critical trials. PE– Prediction Error.

#### 5.3 Data Pre-Processing

Sixty participants completed the task. One participant was excluded as they received a phone call during the task, leaving 59 participants for analysis. In the study phase, two participants demonstrated systematic use of illegal responses (i.e., that did not involve a designated response key) involving buttons adjacent to the legal key. These data were therefore recoded as the corresponding legal responses. Later sensitivity analyses confirmed that this recoding did not significantly affect the main results. Other illegal responses (< 1% of the data) were excluded from the analyses.

#### 5.4 Analysing the Effect of Prediction Error Condition on Memory

All models featured random intercepts for participants and images (i.e., specific photographs of scene categories). Analyses for the familiarisation, training, and testing phases also included by-participant random slopes for PE condition.

Performance accuracy in the familiarisation, training, and study phases was analysed to confirm that the scene–word category associations (priors) were learnt effectively, evidencing that the PE manipulation was successful. For familiarisation, training, and non-critical study trials, the PE condition variable coded whether the scene later (in critical study trials) appeared as a high- or low PE trial. Albeit unlikely given the counterbalancing of stimuli, this verified that there was no significant difference in performance between the high- and low PE scenes before the PE manipulation was performed. Each phase was analysed separately using a logistic mixed-effects model to estimate the probability of a correct response. The PE condition variable was centered in each model so that the intercept evaluated whether performance significantly differed from chance (50% correct).

A logistic mixed-effects model examined the effect of PE condition on memory performance in the testing phase. Trials with stimulus pairings that were not correctly predicted in the study phase were excluded (253 trials; 8.8% of trials) as it was assumed that the PE manipulation was not successful in these trials. An identical model was then run on the data, excluding self-reported guesses to ensure that any effect of PE condition on performance was not driven by these trials.

#### 5.5 Supplementary Analyses: Reliance on Priors as a Function of Schizotypy

Logistic mixed models estimated whether schizotypy significantly influenced the effect of PE condition on the probability of producing a correct response in testing. The models featured random intercepts for images and participants and by-participant random slopes for PE condition. Results revealed no significant relationship between any of the schizotypy metrics and the effect of PE condition on memory performance.

### 6 Mooney Disambiguation Task

#### 6.1 Background

The Mooney disambiguation task measures how the disambiguation of two-tone images is facilitated by exposure to the template figures on which they are based. Support for the role of priors in this phenomenon is provided by fMRI research showing that patterns of early visual cortex responses to Mooney images during post-template viewing resemble responses to templates.^23^ Previous research has indicated that reliance on templates in this task confers a greater advantage in ARMS participants and correlates with hallucination-proneness in controls.^24,25^ In contrast, delusion-prone participants may encode priors as higher-level beliefs, which aid perception of image gist but not finer details.^24^

#### 6.2 Paradigm

This task assesses participants’ ability to discriminate between the figure and background of Mooney images. These images initially appear as meaningless two-tone patterns until participants are presented with the coloured templates on which they are based. This (prior) information enables disambiguation of the image contents, and participants’ use of the prior can be measured as the change in discrimination ability (*d*’) from the pre- to post-template phase.

The task was divided into blocks, each featuring a pre-template, template, and post-template phase. In the pre-template phase, participants decided whether a dot superimposed on the Mooney image was located on-figure (a person or animal) or on-background. This was followed by a template phase, which presented the images that were used to generate the previously encountered Mooney images. Finally, the post-template phase presented the same Mooney images that had appeared in the pre-template trials in order to measure the change in performance due to the use of prior knowledge.

In the pre- and post-template phases, trials began with the appearance of a Mooney image, presented for 1.5 s. A red dot was superimposed on the image 0.2 s after stimulus onset. The image was then replaced with a reminder of the instructions, which asked participants to decide whether the dot had appeared on the figure (left arrow key) or background (right arrow key). After 3 s, participants received an on-screen reminder to respond. The dot appeared equidistant from the centre of the image in all trials to avoid a bias to respond ‘on-background’ for dots placed peripherally. In the template phase, participants viewed coloured templates one by one. Participants were able to click on the template to proceed to the next trial 3 s after stimulus onset. There was no time limit for viewing the templates.

The 5-min training procedure consisted of one block of four trials and served to present all participants with the same instructions and practice stimuli to ensure that they understood the task. This training aimed to demonstrate that Mooney images initially appear as abstract, two-tone stimuli until further information is provided by the templates. The (approximately 25-min) main task featured 24 randomly selected images from the 30 available. Stimuli were presented across eight blocks, with each block testing performance with three images. The pre- and post-template phases used four versions of the three images (12 trials in total), which represented all combinations of the dot placed a) on-figure or on-background and b) on the black or white areas of the image. The template phase in each block repeated the three templates three times (nine trials).

In line with previous research,^24,25^ we hypothesised that there would be a greater *d’* in the post-template phase compared to the pre-template phase.

#### 6.3 Data Pre-Processing

Of the 72 participants who completed the task, three were excluded as they had previously completed a version of the Mooney task and may therefore have had prior experience with the stimuli. A further two participants were excluded because they either misunderstood the instructions or encountered a computer error, leaving 67 participants for analysis.

Only legal responses (involving one of the designated response keys) were analysed. This led to the exclusion of one pre-template trial. Slow (> 5 s) RTs (10 pre-template, 23 post-template trials) were excluded. These could have been caused by a lapse in concentration, forgetting the response keys, or attempting to recall the stimulus to decide on the response. The 5-s threshold was selected to decrease the latter effect of working memory on the task while enabling sufficient time for participants to re-read the response instructions. This fairly liberal cut-off is reasonable as responses were unspeeded.

#### 6.4 Analysing the Effect of Template Exposure on Mooney Disambiguation

The signal detection theory parameter *d*’ was calculated for pre- and post-template phases separately. For all analyses, categorical predictors were dummy coded. To assess the effect of template exposure on figure-ground discrimination, a linear mixed-effects model estimated the effect of (pre- vs. post-template) phase on *d*’, with random intercepts for participants. As there were no time constraints on viewing the templates, a logistic mixed-effects model assessed the effect of template viewing time on post-template phase performance accuracy (percentage correct) and included random intercepts for both participant and image variables. This analysis confirmed that viewing time was unrelated to post-template phase performance.

#### 6.5 Supplementary Analyses: Reliance on Priors as a Function of Schizotypy

Linear mixed models estimated whether each schizotypy metric significantly influenced the change in *d’* from pre- to post-template phase. These included a random intercept for participants. There was no significant effect of any of the schizotypy metrics.

### 7 Gabor Detection Task

#### 7.1 Background

The Gabor detection task^26^ investigates the influence of higher-level beliefs on reports of perceiving near-threshold visual stimuli. In this paradigm, response bias shifts to align with prior information about the likelihood of stimulus presence.^26–28^ Greater prior-responsive shifts in response bias have previously been reported in hallucination-prone participants completing a voice detection task,^29^ suggesting a greater dependence on priors in this group. However, this effect was restricted to priors indicating voice presence, hence hallucination-prone participants may only over-weight priors aligning with their pre-existing perceptual tendencies.

#### 7.2 Paradigm

This task was an adapted version of the paradigm employed by Sherman et al.^27^ and measured the effect of priors on response bias during visual detection. Stimuli were presented using the Psychophysics Toolbox Version 3.^9^

First, a 3-down/1-up staircase was used to compute the contrast of the visual (Gabor) stimulus that would produce a 79% detection rate. This aimed to equate performance across participants so that individual differences in the use of priors in the main task could not be assigned to variations in task difficulty. The Gabor was always present in the lower-right quadrant of the screen in one of two intervals on each trial, with an equal likelihood of appearing in each interval. Participants were instructed to report the interval that featured the Gabor. Each trial began with the central presentation of the text ‘1st’ to indicate the start of the first interval. This was then replaced by a white fixation cross on a grey background followed by a visual search task. The search task was comprised of a circle of four white letters (all either Ls or Ts) that were then quickly masked by white Fs. This procedure (with the same letters) was repeated for the second interval. For the Gabor-present interval, the Gabor was presented at the same time as the circle of letters (before masking). Fifteen percent of trials were catch trials, which also tested participants’ performance on the visual search task (identifying Ls or Ts). The search task ensured that participants fixated centrally instead of directly on the Gabor. Incorrect catch trials were not used to compute the Gabor contrast. The staircase procedure terminated after 10 reversals.

On each trial of the main phase (Figure S3), participants reported the presence/absence of the Gabor in each trial. At the start of each block of trials, participants were presented with (accurate) prior information about the likelihood of Gabor presence in that block. These defined three expectation conditions, with text communicating that the Gabor was ‘likely to be present’ (Gabor-present prior), ‘likely to be absent’ (Gabor-absent prior), or that ‘present and absent trials are equally likely’. These conditions corresponded to 75%, 25%, and 50% probability of Gabor presence, respectively. Reliance on the priors was parameterised by comparing changes in response bias for the 75% or 25% conditions relative to the control (50%) condition. We hypothesised that the Gabor-absent and Gabor-present priors would be associated with a greater (conservative) and lower (liberalised) response bias, respectively, when compared to the control condition.

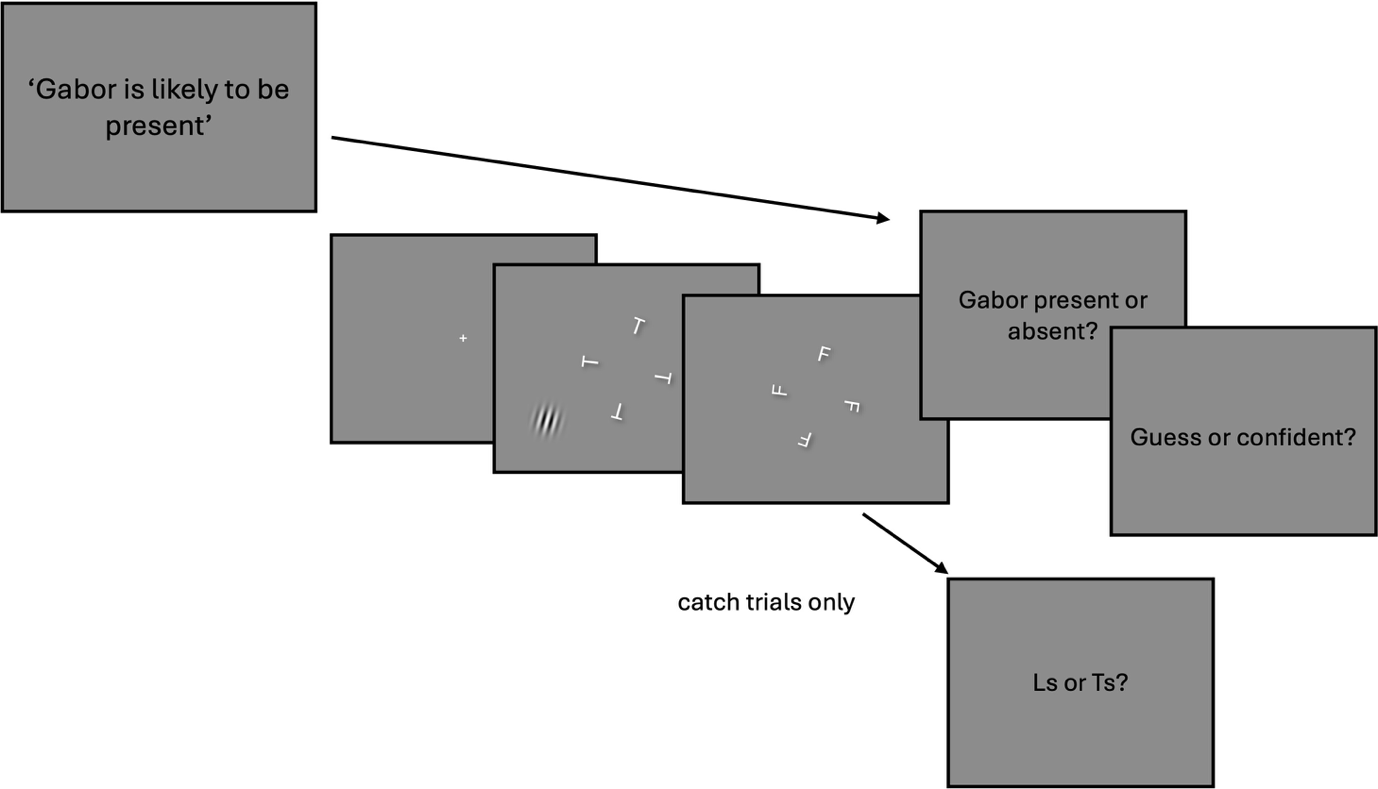

**Figure S3. The Gabor Detection Task.** In each trial, participants reported whether the Gabor stimulus was present (in the lower-right quadrant; shown in the left quadrant in this figure for ease of demonstration). Accurate information about the likelihood of Gabor occurrence (the prior) was presented at the start of each block. On catch trials, participants also reported whether Ls or Ts were present in the visual search task.

#### 7.3 Data Pre-Processing

Sixty-five participants completed the task. One participant was excluded from all analyses as they did not observe the Gabor in any trial. The remaining 64 participants were entered into the analyses.

Analyses of performance on the visual search task confirmed that overall participants performed significantly above chance and accuracy did not vary between the prior conditions. Sensitivity analyses confirmed that removing four participants who demonstrated below-chance visual search performance did not affect the results.

#### 7.4 Analysing the Effect of Priors on Response Bias

Response bias was calculated separately for each prior condition. A linear mixed model investigated whether the priors significantly affected response bias and included random intercepts for participants. The fixed effect of prior was simple coded to create two contrasts comparing the 50% with the 25% and 75% conditions, respectively. The intercept represented the grand mean of the dependent variable (i.e., the mean across all conditions), effectively centering this predictor.^13^ The 50% condition formed the control as in this condition the Gabor stimulus was presented at chance level: there was no informative prior. Analyses of *d’* indicated that discrimination performance improved with higher Gabor contrasts. We therefore included Gabor contrast in the response bias model and confirmed no expectation*Gabor contrast interaction. Furthermore, to assess whether reliance on the Gabor-present prior correlated with reliance on the Gabor-absent prior, we computed a Pearson’s correlation between response bias for these prior conditions.

#### 7.5 Supplementary Analyses: Reliance on Priors as a Function of Schizotypy

Linear mixed models estimated the interaction between schizotypy metrics and the influence of priors on the criterion. These models included random intercepts for participants. Second-level models controlled for Gabor contrast by including this as a fixed effect. Results showed that a higher total PDI score was associated with a reduced reliance on the Gabor-absent prior (*b* = -0.05, *SE* = 0.02, *t* = -2.55, *p* = .012).

### 8 Modelling Schizotypy as a Function of All Task Outcomes

Each schizotypy metric was modelled as a function of all task outcomes. Full model results are shown in Tables S2–6.

|  | ***B*** | **ß** | ***SE*** | ***t*** | ***p*** |
| --- | --- | --- | --- | --- | --- |
| **Intercept** | 18.72 |  | 9.31 | 2.01 | .054 |
| **PP: Perceptual** | -6.00 | -0.14 | 7.76 | -0.77 | .446 |
| **PP: Cognitive** | 4.76 | 0.09 | 10.36 | 0.46 | .649 |
| **Memory** | 0.81 | 0.01 | 16.78 | 0.05 | .962 |
| **Mooney** | 1.30 | 0.07 | 3.60 | 0.36 | .720 |
| **Gabor-Absent Prior** | -5.53 | -0.21 | 6.39 | -0.87 | .394 |
| **Gabor-Present Prior** | 0.46 | 0.01 | 7.54 | 0.06 | .952 |
| **Low Cloze** | 10.98 | 0.10 | 22.44 | 0.49 | .628 |
| **High Cloze** | 14.44 | 0.10 | 26.92 | 0.54 | .596 |
| **CH: No-Tone** | 35.56 | 0.22 | 31.41 | 1.13 | .267 |
| **CH: Subthreshold** | -7.89 | -0.10 | 16.65 | -0.47 | .639 |

**Multiple *R*^2^** = .11

**Adjusted *R*^2^** = -.20

***F*(10,29) = 0.36*, p* = .955**

**Table S2.** **Results From Model of SPQ as a Function of All 10 Measures of Reliance on Priors.** ß represents the standardised coefficient. CH – conditioned hallucinations task; PP– phoneme perception task.

|  | ***B*** | **ß** | ***SE*** | ***t*** | ***p*** |
| --- | --- | --- | --- | --- | --- |
| **Intercept** | 4.67 |  | 2.73 | 1.71 | .098 |
| **PP: Perceptual** | -1.98 | -0.16 | 2.27 | -0.87 | .390 |
| **PP: Cognitive** | -1.38 | -0.09 | 3.04 | -0.46 | .652 |
| **Memory** | -1.42 | -0.06 | 4.92 | -0.29 | .775 |
| **Mooney** | -0.81 | -0.14 | 1.05 | -0.77 | .447 |
| **Gabor-Absent Prior** | -1.83 | -0.23 | 1.87 | -0.98 | .335 |
| **Gabor-Present Prior** | -2.19 | -0.22 | 2.21 | -0.99 | .329 |
| **Low Cloze** | 0.15 | 0.004 | 6.58 | 0.02 | .981 |
| **High Cloze** | 6.85 | 0.16 | 7.89 | 0.87 | .392 |
| **CH: No-Tone** | 0.45 | 0.01 | 9.20 | 0.05 | .961 |
| **CH: Subthreshold** | 2.91 | 0.12 | 4.88 | 0.60 | .555 |

**Multiple *R*^2^** = .14

**Adjusted *R*^2^** = -.15

***F*(10,29) = 0.49*, p* = .885**

**Table S3.** **Results From Model of CAPS as a Function of All 10 Measures of Reliance on Priors.** ß represents the standardised coefficient. CH – conditioned hallucinations task; PP– phoneme perception task.

|  | ***B*** | **ß** | ***SE*** | ***t*** | ***p*** |
| --- | --- | --- | --- | --- | --- |
| **Intercept** | 3.57 |  | 2.72 | 1.32 | .198 |
| **PP: Perceptual** | 0.15 | 0.01 | 2.26 | 0.07 | .946 |
| **PP: Cognitive** | -0.55 | -0.03 | 3.02 | -0.18 | .857 |
| **Memory** | -3.99 | -0.16 | 4.89 | -0.82 | .421 |
| **Mooney** | 0.47 | 0.08 | 1.05 | 0.44 | .660 |
| **Gabor-Absent Prior** | -1.39 | -0.17 | 1.86 | -0.74 | .463 |
| **Gabor-Present Prior** | -3.01 | -0.30 | 2.20 | -1.37 | .182 |
| **Low Cloze** | -4.72 | -0.13 | 6.55 | -0.72 | .477 |
| **High Cloze** | 4.20 | 0.10 | 7.85 | 0.54 | .597 |
| **CH: No-Tone** | 5.33 | 0.11 | 9.16 | 0.58 | .565 |
| **CH: Subthreshold** | 6.12 | 0.25 | 4.86 | 1.26 | .218 |

**Multiple *R*^2^** = .19

**Adjusted *R*^2^** = -.09

***F*(10,29) = 0.68*, p* = .737**

**Table S4.** **Results From Model of CAPS Severity as a Function of All 10 Measures of Reliance on Priors.** ß represents the standardised coefficient. CH – conditioned hallucinations task; PP– phoneme perception task.

|  | ***B*** | **ß** | ***SE*** | ***t*** | ***p*** |
| --- | --- | --- | --- | --- | --- |
| **Intercept** | 5.42 |  | 1.75 | 3.09 | .004* |
| **PP: Perceptual** | -2.51 | -0.28 | 1.46 | -1.72 | .096 |
| **PP: Cognitive** | 0.50 | 0.04 | 1.95 | 0.26 | .800 |
| **Memory** | 0.85 | 0.05 | 3.16 | 0.27 | .790 |
| **Mooney** | -1.07 | -0.26 | 0.68 | -1.58 | .125 |
| **Gabor-Absent Prior** | -2.49 | -0.44 | 1.20 | -2.07 | .048* |
| **Gabor-Present Prior** | 0.34 | 0.05 | 1.42 | 0.24 | .815 |
| **Low Cloze** | 4.47 | 0.18 | 4.23 | 1.06 | .299 |
| **High Cloze** | -4.37 | -0.15 | 5.07 | -0.86 | .396 |
| **CH: No-Tone** | 1.76 | 0.05 | 5.92 | 0.30 | .768 |
| **CH: Subthreshold** | -0.78 | -0.05 | 3.14 | -0.25 | .806 |

**Multiple *R*^2^** = .32

**Adjusted *R*^2^** = .09

***F*(10,29) = 1.38*, p* = .239**

**Table S5.** **Results From Model of PDI as a Function of All 10 Measures of Reliance on Priors.** ß represents the standardised coefficient. CH – conditioned hallucinations task; PP– phoneme perception task.

|  | ***B*** | **ß** | ***SE*** | ***t*** | ***p*** |
| --- | --- | --- | --- | --- | --- |
| **Intercept** | 4.85 |  | 2.26 | 2.15 | .040 * |
| **PP: Perceptual** | -4.64 | -0.33 | 1.88 | -2.46 | .020 * |
| **PP: Cognitive** | 0.67 | 0.04 | 2.52 | 0.27 | .792 |
| **Memory** | -8.40 | -0.31 | 4.07 | -2.06 | .048 * |
| **Mooney** | 0.06 | 0.01 | 0.87 | 0.07 | .947 |
| **Gabor-Absent Prior** | -0.61 | -0.07 | 1.55 | -0.39 | .698 |
| **Gabor-Present Prior** | 3.48 | 0.31 | 1.83 | 1.90 | .067 |
| **Low Cloze** | 11.15 | 0.29 | 5.45 | 2.05 | .0498 * |
| **High Cloze** | -8.18 | -0.17 | 6.53 | -1.25 | .221 |
| **CH: No-Tone** | 17.63 | 0.32 | 7.62 | 2.31 | .028 * |
| **CH: Subthreshold** | 1.16 | 0.04 | 4.04 | 0.29 | .776 |

**Multiple *R*^2^** = .54

**Adjusted *R*^2^** = .38

***F*(10,29) = 3.35*, p* = .005**

**Table S6.** **Results From Model of PDI Severity as a Function of All 10 Measures of Reliance on Priors.** ß represents the standardised coefficient. CH – conditioned hallucinations task; PP– phoneme perception task.

### 9 Distribution of Schizotypy Questionnaire Scores

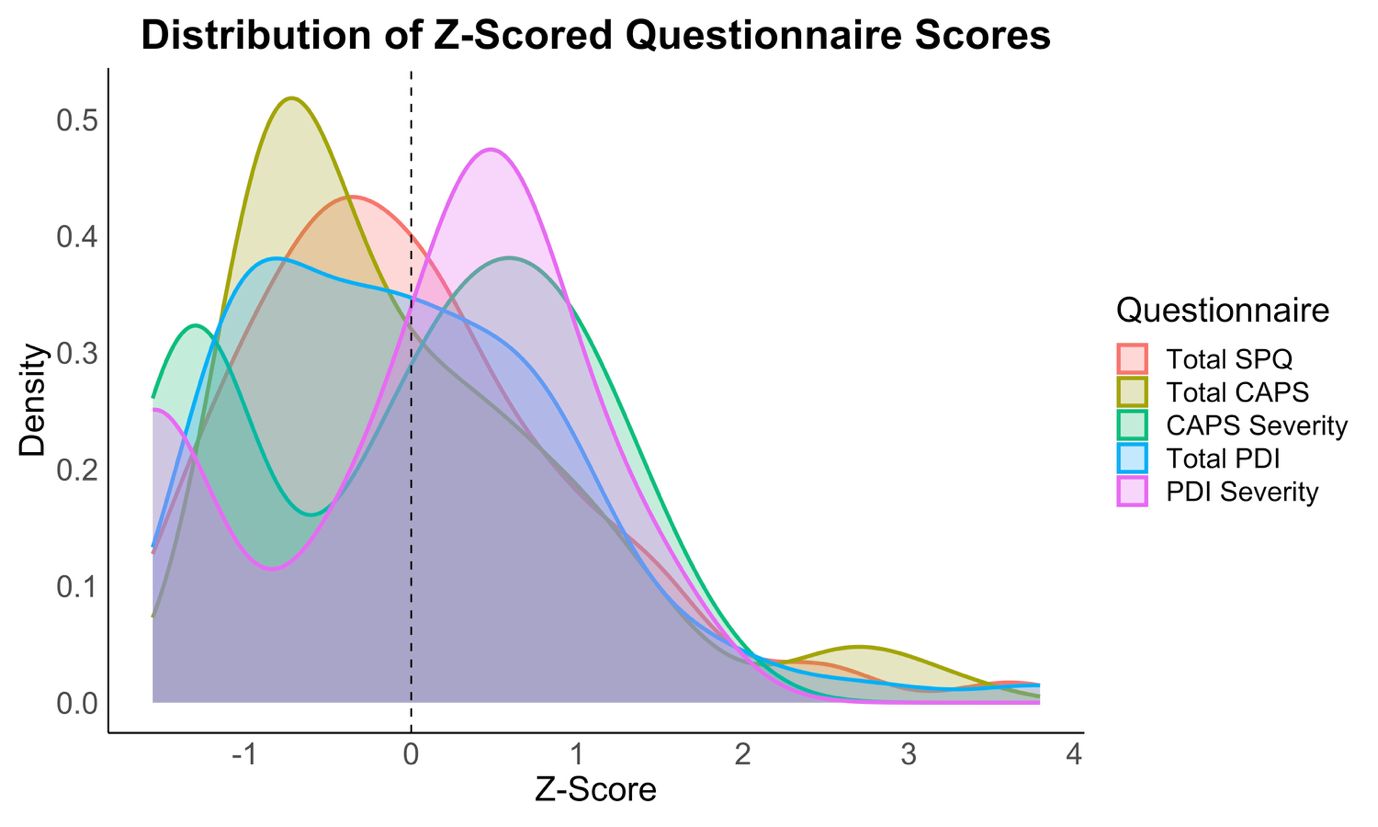

**Figure S4.** **Distribution of *Z*-Scored Questionnaire Scores.** PDI – Peters et al. Delusions Inventory (21-item version)^30^**;** CAPS – Cardiff Anomalous Perceptions Scale^31^; SPQ – Schizotypal Personality Questionnaire.^32^
